# Cerebrospinal fluid concentration of complement component 4A is increased in first-episode schizophrenia

**DOI:** 10.1101/2021.08.17.21262131

**Authors:** Jessica Gracias, Funda Orhan, Elin Hörbeck, Jessica Holmén-Larsson, Neda Khanlarkani, Susmita Malwade, Sravan K. Goparaju, Lilly Schwieler, Helena Fatourus-Bergman, Aurimantas Pelanis, Carleton P. Goold, Anneli Goulding, Kristina Annerbrink, Anniella Isgren, Timea Sparding, Martin Schalling, Viviana A. Carcamo Yañez, Jens C. Göpfert, Johanna Nilsson, Ann Brinkmalm, Kaj Blennow, Henrik Zetterberg, Göran Engberg, Fredrik Piehl, Steven D. Sheridan, Roy H. Perlis, Simon Cervenka, Sophie Erhardt, Mikael Landen, Carl M. Sellgren

## Abstract

Excessive synapse loss is a core feature of schizophrenia and is linked to the complement component 4A gene (*C4A*). In two independent cohorts, we show that cerebrospinal fluid (CSF) C4A concentration is elevated in first-episode psychosis patients who develop schizophrenia and correlates with CSF measurements of synapse density. Using patient-derived cellular modeling, we find that disease-associated cytokines increase neuronal *C4A* expression and that IL-1β associates with C4A in patient-derived CSF.

## Main Text

Schizophrenia (SCZ) is a highly heritable and polygenic brain disorder with the strongest associated SCZ locus located close to the complement component 4 (*C4*) genes. Distinct from mouse, human *C4* is encoded by two closely related genes, *C4A* and *C4B*, typically present in multiple copy numbers (CNs) per genome and gene isotype. SCZ risk attributed to this locus can largely be explained by genetically predicted *C4A* RNA expression, and elevated *C4A* expression has been confirmed in SCZ postmortem brain tissue^1^. During brain development, microglia utilize complement signaling for selective removal of supernumerary synapses by complement receptor 3 (C3R)-dependent phagocytosis^2,3^. In line with the observed decrease in synapse density in SCZ^4,5^, excessive microglial synapse elimination has been observed in SCZ-derived in vitro models^6^. This suggests that the increased SCZ risk in microglial synapse removal in early stages of the disease. *C4A* CNs also correlate with complement deposition in patient-derived models, as well as microglial engulfment of synaptic structures^6^. Notably, *C4B* CNs, not linked to SCZ risk^1^, do not influence neuronal complement deposition or synapse elimination in these models^6^. Recently, these in vitro findings were confirmed in vivo using mouse models overexpressing human *C4A* or *C4B*^7^.

Despite the accumulating evidence from experimental models linking *C4A* to excessive synapse removal in SCZ, data showing increased in vivo protein levels in patients has been lacking. One reason is that these measurements have been limited by difficulties in distinguishing C4A and C4B as the peptide sequence only differ by a few amino acids. In the present study, we developed a targeted mass spectrometry method capable of detecting unique peptide sequences in C4A and C4B protein. We applied this method to human cerebrospinal fluid (CSF) collected from two independent cohorts of first-episode psychosis (FEP) patients and healthy controls (HCs) (**Figure 1a**).

**Figure 1.**
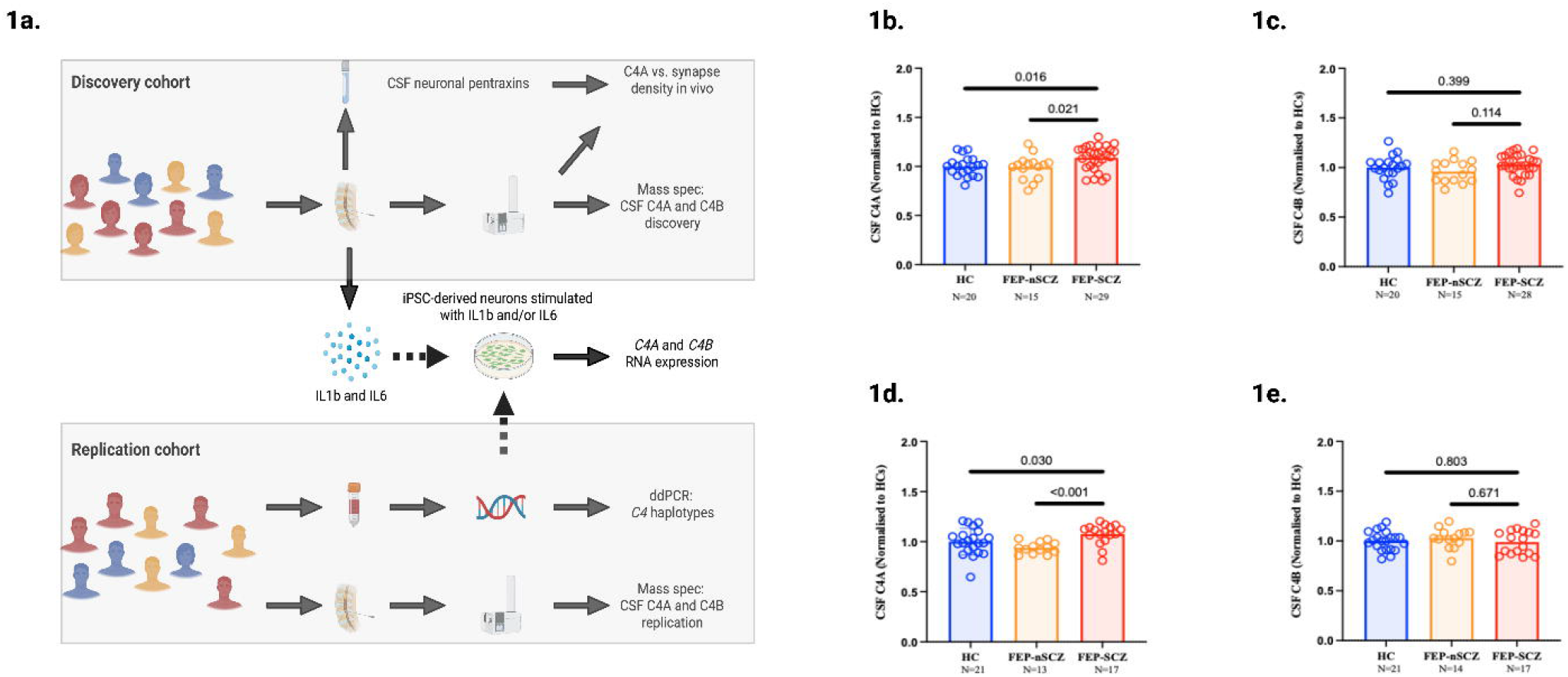
Cerebrospinal fluid levels of C4A is increased in first-episode psychosis patients who develop schizophrenia. **a**. Overview of the study design. **b**. In the discovery cohort (KaSP), first-episode psychosis patients who developed schizophrenia (FEP-SCZ) displayed significantly higher cerebrospinal fluid (CSF) C4A concentrations as compared to healthy controls (HCs) or FEP patients who did not develop SCZ (FEP-nSCZ). **c**. CSF C4B concentrations were similar across groups. **d**. In the replication cohort (GRIP), FEP-SCZ patients displayed significantly higher CSF C4A concentrations as compared to HCs or FEP-nSCZ patients, while (**e**.) CSF C4B concentration were similar across groups. Combining the both cohorts we observed higher CSF C4A concentrations in FEP-SCZ patients as compared to both FEP-nSCZ and HCs (**Supplementary Figure 6**). Bar graphs represent means (with CSF C4A concentrations normalized to the mean of HCs). Data were analysed using Brown-Forsythe and Welch analysis of variance (ANOVA) models and adjusted for multiple testing. Significance was set to *P*<0.05. CSF C4A and C4B concentrations were log transformed before analysed. All reported p-values are two-sided.

In the first cohort (KaSP; discovery sample) we included 44 FEP patients and 20 age- and sex-matched HCs. FEP patients were stratified depending on if they subsequently developed SCZ (FEP-SCZ; 29 subjects) or not (FEP-nSCZ; 15 subjects). The groups displayed no significant differences in terms of demographics and clinical characteristics (**Supplementary Table 1**). We observed significantly higher CSF C4A concentrations in FEP-SCZ patients as compared to HCs or FEP-nSCZ patients (**Figure 1b**). On the contrary, CSF C4B concentrations were similar across the three groups (**Figure 1c**). Fourteen FEP-SCZ patients and 8 FEP-nSCZ patients had been prescribed an antipsychotic, although in no instance for more than one month before CSF collection. At the group level, concentrations of both C4A and C4B were similar between patients who had been exposed to an antipsychotic and those being antipsychotic-naïve, and FEP-SCZ patients displayed higher C4A concentrations also after adjusting for the use of antipsychotic medication (**Supplementary Figure 1**).

Results were validated in an independent FEP cohort (GRIP; replication sample), comprising of 31 FEP patients (FEP-SCZ; n=17, FEP-nSCZ; n=14) together with 21 HCs. FEP-nSCZ patients were more commonly smokers than HCs but otherwise the three groups did not differ in terms of demographics and clinical characteristics (**Supplementary Table 2**). However, smoking was unrelated to CSF C4A or C4B concentration (*r*_*pearson(p)*_=−0.16; *P*=0.253, and *r*_*(p)*_=−0.23; *P*=0.102, respectively). In accordance with our results from the discovery cohort, we observed significantly higher CSF C4A concentrations in the FEP-SCZ group as compared to HCs or the FEP-nSCZ group (**Figure 1d**), while CSF C4B concentrations were similar across groups (**Figure 1e**). Twenty-two patients (9 FEP-SCZ and 13 FEP-nSCZ) were prescribed an antipsychotic, with FEP-SCZ patients still displaying higher CSF C4A concentrations compared to FEP-nSCZ patients after adjusting for use of antipsychotic medication (**Supplementary Figure 2**).

Clinical data regarding severity and symptom profiles were available for the KaSP cohort (n=44), and were used for exploratory analyses of correlations between CSF C4A concentrations and disease phenotypes. After controlling for multiple testing, we observed no significant correlations between CSF C4A concentration and scores on the positive, negative, and general psychopathology scale of the Positive and Negative Syndrome Scale (PANSS), or performance in the key domains of the MATRICS Consensus Cognitive Battery (MCCB) (**Supplementary Table 3**). We then studied CSF C4A in the context of SCZ-related impairment of sensorimotor gating by measuring pre-pulse inhibition (PPI) and habituation due to repeated exposure to a stimulus^8,9^. Similar to findings in mouse studies, CSF C4A displayed no significant association with % PPI at either interstimulus interval^7^. However, we observed an association with decreased habituation (**Supplementary Table 4**), a deficit hypothesized to reflect impairment in synaptic plasticity^10^.

Despite a strong correlation between *C4A* CNs and *C4A* RNA expression in the brain^1^, previous studies suggest that upregulation of *C4A* expression in SCZ exceeds what can be expected from genetic risk variance in the *C4* locus^11^. To test this at a protein level, we used droplet digital PCR (ddPCR) to measure *C4A* CNs and *HERV* insertions in FEP patients with available DNA and CSF C4A protein levels (n=22 FEP and n=21 HCs; GRIP cohort). We observed a robust correlation between *C4A* CNs and CSF C4A protein levels (*r*_*spearman(s)*_=0.47; *P=*0.002), as well as between genetically predicted *C4A* RNA expression^1^ and CSF C4A protein levels (*r*_*(s)*_=0.44; *P=*0.003), while CSF C4A levels adjusted for *C4A* CNs were still higher in FEP-SCZ patients as compared to FEP-nSCZ patients and HCs (**Supplementary Figure 3**). A recent *C4A*-seeded brain co-expression network analysis has implicated an enrichment of genes involved in immune processes, such as NF-κB signaling, rather than other complement system genes^11^, and experimental studies suggest that cytokines can modulate the expression of the *C4* genes although in a tissue-dependent fashion and with a variable effect on *C4A* versus *C4B* expression^12,13^. The pro-inflammatory cytokines, interleukin(IL)-1β and IL-6 both have repeatedly been shown to be elevated in CSF of SCZ patients^14,15^, and we therefore explored their influence on neuronal *C4A* expression. To this end, we generated stably inducible *neurogenin 2* (*NGN2*) neural progenitor cells from an SCZ patient-derived induced pluripotent stem cell line, with equal CNs of *C4A* and *C4B*, and differentiated these cells to cortical excitatory neurons. These cultures were stimulated for 24 hours with either IL-1β (1ng/ml), IL-6 (1ng/ml), or a combination of IL-1β and IL-6 (**Figure 2a**). We observed significantly increased expression of *C4A* as measured by qPCR when IL-1β and IL-6 were combined (**Figure 2b**), while no such effect was evident for expression of *C4B* (**Figure 2c**), thus suggesting that the induction is predominately influencing *C4A* expression. To corroborate this observation in a clinical context and to determine the relative importance of each cytokine in clinically relevant concentrations, we measured CSF IL-1β and IL-6 in 25 FEP patients with available CSF (KaSP cohort). Controlling for genetically predicted *C4A* RNA expression using available whole genome sequencing data^1^, we observed a significant positive correlation between IL-1β and CSF C4A concentration (**Figure 2e**), while the correlation between IL-6 and CSF C4A concentration was less pronounced and did not reach significance (**Figure 2f**). In contrast, there was no correlation between these two cytokines and CSF C4B concentrations (**Figure 2g** and **2h)**. Collectively, this suggests that in FEP patients primarily elevated IL-1β levels could contribute to excessive synaptic pruning by specifically inducing *C4A* expression per *C4A* CN.

**Figure 2.**
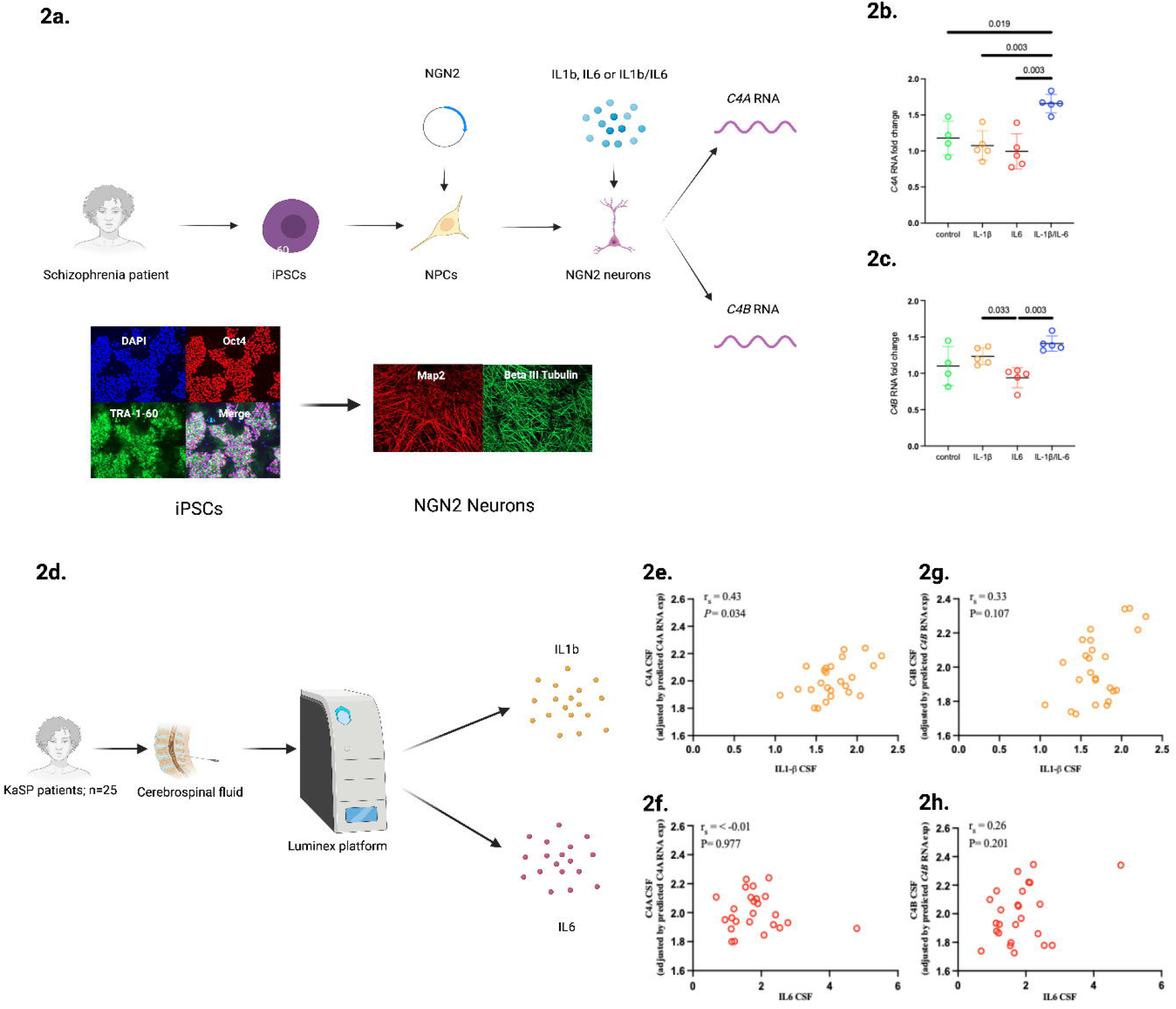
Disease-associated cytokines increase neuronal *C4A* expression in a patient-derived schizophrenia model and associate with C4A protein levels in cerebrospinal fluid from first-episode psychosis patients. **a**. Overview of in vitro experiments. Induced pluripotent stem cells (iPSCs) derived from a schizophrenia patient with two copy numbers of *C4A* and *C4B*, respectively. Quality control immunocytochemistry images showing octamer-binding transcription factor 4 (POU domain, class 5, transcription factor 1) and TRA-1–60, as well as nuclear staining with DAPI for iPSCs that were used to derive cortical excitatory neurons (stained for MAP2 and BetaIIItubulin). **b**. *C4A* RNA expression increased in cortical excitatory neurons after pre-treatment for 24 hours with interleukin(IL)-1β and IL-6, while **c**. no significant increase was observed for *C4B* RNA expression. **d**. Overview of in vivo analyses. IL-1β, but not IL-6, displayed a significant association with C4A protein levels in cerebrospinal fluid (CSF, adjusted for genetically predicted *C4A* RNA expression), while none of the cytokines displayed significant associations with CSF C4B protein levels. **e**. - **f**. display the correlations (Spearman’s correlation coefficient) in separate models for CSF C4A, and in **g**.-**h**. for CSF C4B. Data in b. and c. were analyzed using Brown-Forsythe and Welch analysis of variance (ANOVA) models and adjusted for multiple testing. Corrected p-values were then compared to a 0.05/2 significance threshold given analyses of both *C4A* and *C4B*. qPCR was performed in three independent experiments, each data point is an average of triplicate values and CSF was analysed from 25 patients. All p-values are two-sided and bar graphs represent means.

Lastly, as *C4A* CNs predict synaptic complement deposition and synaptic pruning in experimental models^6^, we explored associations between CSF C4A protein concentrations and an in vivo proxy of synapse density. CSF levels of neuronal pentraxins (1, 2, and receptor) were recently shown to inversely correlate with prefrontal cortical thickness determined with MRI, as well as to predict synapse loss in dementia and Alzheimer’s disease^16,17^. Thus, we measured these three markers in CSF from the KaSP cohort (FEP patients n=43; HCs n=19). After adjusting for multiple testing, we observed significant negative correlations between C4A and neuronal pentraxin 1 and 2, as well as the neuronal pentraxin receptor **(Supplementary Table 5)**.

In conclusion, this study demonstates that C4A protein concentrations are elevated in the CSF of FEP patients who subsequently develop SCZ, strenghtening the notion of a role for this complement component in the disease pathogenesis. Furthermore, in line with the effect of *C4A* CNs on microglial synapse engulfment in patient-derived models, we show that C4A protein levels in patient-derived CSF inversely correlate with proxies of synapse density. We also corroborate previous results that *C4A* CNs do not fully explain the observed *C4A* RNA upregulation in SCZ^1,11^, as *C4A* CNs cannot fully explain the increased CSF C4A protein levels in FEP-SCZ patients seen here. This aligns with our findings that SCZ-related cytokines can induce neuronal *C4A* expression in a patient-derived experimental model and that CSF IL-1β concentrations correlate with CSF C4A levels adjusted for genetic variance in the *C4* locus.

## Methods

### Ethical statement

The study was approved by the Regional Ethics Committee in Stockholm (Sweden) and the Institutional Review Board of Partners HealthCare (MA, USA). Informed consent was obtained from all included subjects.

### Study populations and diagnostic assessments

KaSP Cohort: Karolinska Schizophrenia Project (KaSP) is a Swedish multidisciplinary research consortium investigating the pathophysiology of FEP and SCZ. Patients who seek health care for psychotic symptoms for the first time are recruited at psychiatric emergency wards and in- or outpatient facilities at the psychiatric clinics located in Stockholm, Sweden. Exclusion criteria are ongoing or previous prescription of an antipsychotic for more than 30 days, severe somatic and neurological diseases, current substance abuse (except nicotine use), or autism spectrum disorder. These patients are excluded by clinical examination, medical history, routine laboratory tests, including screening for drugs and MRI scans. A baseline diagnosis is established based on a structured clinical interview of the Diagnostic and Statistical Manual of Mental Disorders IV, DSM-IV (Structured Clinical Interview for DSM-IV-Axis I Disorders, SCID-I) at the time of inclusion. For the subdivision of FEP patients into a FEP-SCZ and a FEP-nSCZ group we used data from a 1.5-year follow-up (median time to follow-up: 1.6 years). The same diagnostic procedure was used as at baseline. Patients who did not receive a SCZ diagnosis at follow-up received a diagnosis of delusional disorder, unspecified psychosis, acute and transient psychotic disorder, schizoaffective disorder, or major depressive disorder. HCs were recruited by advertisement and matched on age and sex. Eligibility was determined by medical history, routine laboratory tests, clinical examination, and an MRI examination, as evaluated by an experienced neuroradiologist at the MR Centre, Karolinska University Hospital, Solna. The Mini International Neuropsychiatric Interview (MINI), performed by either a resident or a specialist in psychiatry, was used to exclude previous or current psychiatric illness. Further exclusion criteria were former or current use of illegal drugs, first-degree relatives with psychotic illness or bipolar disorder, as well as neurologic disease and/or severe somatic disease. All participants (patients and controls) were free from any form of substance abuse evaluated with Alcohol Use Disorders Identification Test (AUDIT) and the Drug Use Disorders Identification Test (DUDIT) at the time of the study.

GRIP cohort:The Gothenburg research and investigation on psychosis (GRIP) is a study collecting data from patiens with schizophrenia spectrum disorders at the psychosis clinic, Sahlgrenska University Hospital, Mölndal, Sweden. The initial diagnostic assessments are performed by treating clinicians at outpatient tertiary-care units, as well as the inpatient units, in Sahlgrenska University Hospital (Sweden). Further clinical information is obtained from interviews at tertiary-care units and investigation units, and when needed supplemented with information from clinical records. To secure diagnoses for research, a series of case conferences are then held (for this study in between October 2017 – March 2018). A minimum of two board-certified psychiatrists participate in these case conferences where a consensus diagnosis is established according to the Diagnostic and Statistical Manual of Mental Disorders, Fifth Edition (DSM-5). The diagnosis is based on structured interviews, medical records (also collected post-baseline), and when needed supplemented with information from the treating psychiatrist. The participating research clinicians are blinded to the results of the CSF analyses. The median time from inclusion in the study (when the initial diagnosis by the treating physician is made) to the establishment of a research consensus diagnosis on the case conference was here 1.8 years. For FEP subjects, the median time from first contact with psychiatric care to referral for further investigation of psychotic symptoms at the tertiary unit (i.e., study baseline) was 2 years. This time period, as well as time to SCZ diagnosis within the study period, was unrelated to CSF C4A or C4B levels, as well as to a SCZ-vs. a non-SCZ-related diagnosis (data not shown). Patients who did not receive a SCZ diagnosis at the diagnosis evaluation around 2 years after baseline received a diagnosis of bipolar disorder, brief psychotic disorder, substance-induced psychotic disorder (not known at the initial evaluation), or a psychotic disorder not otherwise specified. As no matched HCs are recruited in GRIP we used HCs from the larger “St. Göran projektet” research project in which GRIP is a sub-study focusing on psychosis. These HCs are randomly selected by Statistics Sweden and undergo the same clinical investigations as patients and do not fulfil the criteria for a DSM-IV-TR disorder. From this cohort, we then selected HCs to as closely as possible match the patients in GRIP dependent on sex and age.

### Lumbar puncture and collection of CSF

KaSP cohort: Study participants fasssted overnight, lumbar punctures were performed between 7.45 am – 15.15 pm, using a non-cutting 22G spinal needle inserted at the L4–5 interspace with collection of 18 mL of CSF. Not more than 1 h after collection, CSF was centrifuged at 3500 rpm for 10 minutes divided into 10 aliquots and stored at –80°C until analysis.

GRIP cohort: Subjects fasted overnight, and the lumbar puncture was performed between 10.45 and 11.15 a.m. A non-cutting 22G spinal needle was inserted into the L4/L5 interspace and about 15 ml of CSF was collected, gently inverted to avoid gradient effects, centrifuged for 10 minutes at 3500rpm at 20 degrees, not more than 30 minutes after sampling. CSF was then divided into 0.5 ml aliquots that were stored at −80°C pending analysis.

### Targeted Mass Spectrometry (C4A and C4B)

The protein concentration of the CSF samples was measured in triplicates using Micro BCA Protein Assay Kit (Thermo Fisher Scientific Inc, USA PN: 23235) prior to sample preparation. The measurements were performed in a 96-well plate (VWR International AB, Sweden PN: 738-0170) and fluorescence read by plate reader FLUOstar Omega. (BMG LABTECH, Germany). Sample preparation was performed on the Agilent AssayMAP Bravo Platform. Fifty µg protein of each CSF sample was used for analysis. Depending on the protein concentration of the samples, the appropriate volume of each sample was transferred to a 96-well plate (Greiner G650201) and 200 mM triethylammonium bicarbonate buffer (TEAB, Thermo Fisher Scientific, USA PN: 90114) manually added to a final volume of 50 µL. The Bravo platform was used for the following steps; addition of 50 µL TEAB buffer and 50 µL 3.3% solution of sodium deoxycholate (SDC, Sigma-Aldrich, Sweden PN: L3771) for dissociation of proteins (final concentration of 1%). Proteins were reduced by addition of 10 µL 80 mM tris(2-carboxyethyl) phosphine (TCEP, final concentration of 5 mM, Sigma-Aldrich, Sweden PN: C4706) and the reaction was carried out for 1 hour at 55°C. Alkylation was carried out by addition of 15 µL 120 mM iodoacetamide (final concentration of 10 mM, Sigma-Aldrich, Sweden PN: I1149) for 30 min in the dark at room temperature. One µg trypsin (Sequencing Grade Modified, Promega, Sweden PN: V5111) was added to each sample and incubated overnight at 37 °C. The CSF samples were divided into four groups for sample preparation and they were labeled with #1, #2, #3 and #4. In group #3, the protein concentration was lower than in the other groups and the starting volume for each sample was therefore more than 50 µL. To these samples, 200 mM triethylammonium bicarbonate buffer was manually added to a total volume of 100 µL and additional liquid handling steps were carried out on the Bravo platform. The trypsin digestion was quenched by addition of 30 µL 10% FA (Thermo Fisher Scientific, USA PN: 94318) and the SDC pellet was removed by filtering the samples through a polypropylene filter plate (Agilent Technologies, Sweden, PN: 200931-100) with hydrophilic PVDF membrane (mean pore size 0.45 µm). The tryptic peptide concentration was detected using the Pierce Quantitative Colorimetric Peptide Assay (Thermo Fisher Scientific, USA PN: 23275). Each sample was spiked with a mixture of heavy isotope-labeled peptide standards (AQUA QuantPro peptides from Thermo Fisher Scientific) before LC-MS analysis.

The LC-MS analysis was performed on a Tribrid mass spectrometer Fusion equipped with a Nanospray Flex ion source, coupled to an EASY-nLC 1000 ultra-high pressure liquid chromatography (UHPLC) system (Thermo Fisher Scientific, USA). The tryptic CSF peptides spiked with AQUA peptides, were loaded on to an Acclaim PepMap 100 C18 precolumn (75 μm x 2 cm, Thermo Fisher Scientific, USA) and separated on an Acclaim PepMap RSLC column (75 μm x 25 cm, nanoViper, C18, 2 μm, 100 Å) with the flow rate 300 nL/min. The column was kept at 40°C using a Phoenix S&T Nano LC column heater (MS Wil GmbH, Zurich). Solvent A (0.1% FA, Sigma-Aldrich) and solvent B (0.1% FA in acetonitrile, Sigma-Aldrich) were used for the nonlinear gradient. The percentage of solvent B was 1% for the first 3 min and increased to 30% in 50 min and then to 90% in 5 min which was kept for 7 min to wash the column. The tryptic CSF peptides spiked with AQUA peptides, were introduced into the mass spectrometer via a stainless-steel Nano-bore emitter (OD 150 µm, ID 30 µm) with the spray voltage of 2 kV and capillary temperature 275 °C. The Orbitrap Fusion was operated in parallel reaction monitoring (PRM) mode. MS2 precursors were isolated with a quadrupole mass filter set to a width of 1.2 m/z. Precursors were fragmented by higher energy collision dissociation (HCD) and detected in Orbitrap detector with a resolution of 60.000. The normalized collision energy (NCE) for HCD was 25%. The values for the AGC target and maximum injection time were 5 × 10^4^ and 118 ms, respectively. The peptide inclusion list is shown in **Supplementary Table 6**. Five µL (0.2 µg/µL) of tryptic peptides (spiked with AQUA peptides) of each trypsin digested CSF sample, was injected and run in triplicates. Pooled CSF samples were run as quality control samples throughout all the samples.

The linear range of each heavy peptide was measured in pooled CSF digest. A mixture of heavy AQUA peptides was spiked at five different concentrations into pooled CSF samples. The concentration range was adjusted for each individual peptide according to its expected endogenous signal. Each sample was analyzed three times, and the peak areas of heavy peptides were plotted against the spiked heavy peptides concentrations (**Supplementary Figure 4**).

All the raw data generated on the Fusion MS were imported to Skyline v4.1 (MacCoss Lab Software, USA) for data analysis. Peak integration was done automatically by the software and was manually inspected to confirm correct peak detection. Peak identities were confirmed by measured transitions (dotp) and also between endogenous and corresponding heavy peptides (rdotp). The best 3 to 5 transitions were selected for the quantification and was performed by matching light and heavy peak area ratios. All calculations were performed using Microsoft Excel.

For quality control, we also measured a peptide shared for C4A and C4B (tC4; GRIP). The sum of C4A + C4B peptide levels in the combined sample strongly correlated to tC4 (*r*_*s*_=0.91; P<1×10^−15^). One subject with *C4A* CNs also displayed non-detectable levels of the C4A peptide. We decided to exclude all subjects not displaying detectable peptide levels in the main analyses. However, analyses were also performed including zero values, as well as imputing zero values with half of the limit of detection, but with very similar results (data not shown).

### Molecular analysis of *C4* structural elements (ddPCR)

*CN*s of *C4* structural elements (*C4A, C4B, C4-HERV* CNs) in the GRIP cohort were measured using ddPCR as previously described ^6^. *C4A-HERV+, C4A-HERV-, C4B-HERV+*, and *C4B-HERV-CN*s were determined based on imputations as previously described^1^. For distributions of *C4 CN*s in the samples see **Supplementary Figure 5**. *C4A*, or *C4B, CN*s displayed no association to age or sex (data not shown).

### Molecular analysis of *C4* structural elements (imputation from WGS)

*CN*s of *C4* structural elements (*C4A-HERV+, C4A-HERV-, C4B-HERV+*, and *C4B-HERV-CN*s) in the KaSP cohort were imputed from MHC genotypes computed from whole genome sequencing (WGS) data. HapMap3 CEU reference haplotype panel, from Sekar et al., 2016^1^, was used to impute pre-annotated C4 haplogroups, using Beagle (version 3.3), with subsets of SNPs in the extended MHC locus (chr6: 25-34 Mb).

### Cognitive testing

The Measurement and Treatment Research to Improve Cognition in Schizophrenia Consensus Cognitive Battery was used to evaluate cognitive function in the KaSP cohort. This battery contains 10 tests that measure 7 cognitive domains: Speed of processing (Brief Assessment of Cognition in Schizophrenia: Symbol Coding, Category Fluency: Animal Naming, Trail Making Test: Part A); Attention/vigilance (Continuous Performance Test-Identical Pairs); Working memory (Wechsler Memory Scale-3rd Edition: Spatial Span, Letter-Number Span); Verbal learning (Hopkins Verbal Learning Test-Revised); Visual learning (Brief Visuospatial Memory Test-Revised); Reasoning and problem solving (Neuropsychological Assessment Battery: Mazes) and Social cognition (Mayer–Salovey–Caruso Emotional Intelligence Test: Managing Emotions). One psychologist (HFB) administered all the tests.

### Sensorimotor gating

Electromyography (EMG) was recorded from the orbicularis oculi using two 20-mm disk Ag/AgCl electrodes positioned below and lateral to the eye. A 40×50 mm ground electrode was implanted behind the ear on the bone. EMG activity was filtered (1- to 1000-Hz notch filter and 60-Hz notch filter) and digitized at a rate of 1 kHz. Acoustic stimuli were supplied via headphones using a Psylab Stand Alone monitor and a tone generator (containing a digital white noise source and a digitally controlled sine wave generator with a range of 30 to 2000 Hz from Contact Precision Instruments). The EMG signal was amplified with a Grass A.C. Amplifier (model 1CP511, Astro-Med., Inc.) and acquired by using commercially available hardware/software (BioPac) on a laptop computer (Hewlet Pacard Compaq 6715b). All subjects (KaSP) were tested using the same startle system. Subjects were instructed to relax and keep their eyes open and directed against a position marked on the wall, during the test. Throughout the session, background noise was set to 70 dB. The session consisted of 60 trials with two conditions: pulse-alone trials, consisting of a 40 ms, 115-dB pulse of white noise and prepulse-pulse trials, consisting of a 40 ms 115 dB pulse (white noise, as above) preceded by a 16-dB (above background) prepulse of 20 ms duration (white noise). The ISI between the prepulse and the pulse tested were 30, 60, and 120 ms, respectively. A 3-min acclimation period with 70 dB white noise started the session. The immediately following block consist of 15 pulse-alone trials, used for calculation of habituation using the latent curve modeling^18^. The next three blocks each consisted of 15 trials: 4–5 pulse-alone trials and 5 prepulse-pulse trials with the ISI of interest and 5 prepulse-pulse trials with other ISIs (not included in the analysis of PPI for the ISI of interest). Processing of the the EMG recording was done offline with a 100-Hz high-pass filter and baseline correction by using a 100 ms prestimulus baseline. Response onset was defined by the first crossing from baseline within a 20–120 ms window after stimulus onset. The difference of the most positive peak and most negative in a 20–150 ms window after pulse onset was used to calculate the peak response amplitude. The following startle measures were examined; 1) reactivity, or the magnitude of response (startle amplitude (SA), 2) PPI or the percentage of change in startle magnitude to prepulse + pulse versus pulse-alone trials (((pulse − prepulse + pulse) / pulse) * 100)^19^, 3) habituation or the decrement in reactivity with repeated stimulus administration. The primary outcome measures of PPI were calculated for 30, 60, and 120 ms ISI conditions, respectively, as percent change in response amplitudes [% PPI = (mean startle alone-mean prepulse-pulse condition)/mean startle alone × 100]. Startle response was calculated by comparing the mean startle magnitude of the middle five pulses during the first trial (that only contained pulse alone).

### iPSC generation and neuronal differentiation

Human fibroblasts from a male with SCZ (2 *CN*s of *C4A* and 2 *CN*s of *C4B*) were reprogrammed, stabilized, and expanded under xeno-free conditions by Cellular Reprogramming, Inc. (www.cellular-reprogramming.com), as previously described previously^6^. Briefly, induced pluripotent stem cell (iPSC) colonies were obtained using mRNA reprogramming in a feeder-free culture system. Stable iPSCs were expanded in NutriStem XF medium (Biological Industries) and on biolaminin 521 LN-coated (BioLamina) plates to at least passage 3. iPSCs were then purified using MACS with anti-TRA-1–60 MicroBeads (Miltenyi Biotec) on LS columns according to the manufacturer’s instructions. All fibroblasts and iPSCs were screened and found negative for Mycoplasma; they stained positive for octamer-binding transcription factor 4 (POU domain, class 5, transcription factor 1) and TRA-1–60 **(Figure 2a)**.

We then generated *NGN2* expressing stable NPC lines using TALEN-based plasmids as previously described^6^. Briefly, the doxycycline-inducible *NGN2* AAVS1 knock-in plasmid, based on an AAVS1 SA-2A-puro-pA donor (plasmid no. 22075; Addgene), was generated by replacing the puromycin resistance gene with the neomycin resistance gene and cloning a cassette containing the Tet-On 3G promoter driving the human *NGN2* complementary DNA followed by P2A, a zeocin resistance gene, the BGH polyA, CMV early enhancer/chicken β - actin (CAG) promoter driving Tet-On 3G and BGH polyA. The AAVS1-targeting TALENs are based on hAAVS1 1R TALEN and hAAVS1 1L TALEN (plasmid nos. 35432 and 35431, respectively; Addgene) and were generated by golden gate assembly. Plasmid maps can be provided upon request. NPCs were collected with Accutase and counted. For each transfection, 4 × 10^6^ cells were pelleted at 300g and then resuspended in 100 μL prewarmed nucleofector solution (Human Stem Cell Kit 1, Lonza, catalog no. VPH-5012); 4 μg of NGN2 plasmid and 1.5 μg each of 1R and 1L plasmids were added directly to the resuspended cells, or negative control without plasmid, followed by nucleofection (Amaxa Nucleofector II; Lonza) according to the manufacturer’s protocol using program B-016. After nucleofection, cells were plated onto Geltrex-coated 6-well plates in neural expansion medium (50% NBM, 50% advanced DMEM/F-12 with 1× NIS). Stable lines were expanded by selection with the addition of 125 μl mL^−1^ G418 (Geneticin; Thermo Fisher Scientific) over 8–10 d or until the negative transfection control cells completely died.

*NGN2* neurons were then generated as described earlier^6^. Briefly, NPCs were plated as single cells on poly-L-ornithine/laminin coated 24-well polypropylene plates at 2.5×105 cells/500 μL in 50% neurobasal medium (NBM; ThermoFisher Scientific) and 50% DMEM/F12 (ThermoFisher Scientific) with 1x neural induction supplement (Thermo Fisher Scientific). On the following day, the medium was replaced with neuronal differentiation medium (50% NBM and 50% DMEM/F12 supplemented with 1x MEM Non-essential Amino Acids (ThermoFisher Scientific), 1x N-2 supplement (ThermoFisher Scientific), 10 ng mL^−1^ brain-derived neurotrophic factor (BDNF; Peprotech, Cat# 450-02) and 10 ng m L^−1^ recombinant human NT-3 (PeproTech, Cat# 450-03)). Day 2 cells were differentiated in neuronal differentiation medium enriched with 1× B-27 (ThermoFisher Scientific) and 2 μg m L^−1^ doxycycline (Sigma-Aldrich, Cat# D3073). This medium was maintained for the entire differentiation process, unless noted. On days 3-6, cells were incubated with the same medium with the addition of 5 μg m L^−1^ zeocin (ThermoFisher Scientific). For neuronal maturation, 11 days after seeding, astrocyte conditioned medium (ACM: ScienCell Research Laboratories, Cat# SC1811-sf) was added to the culture. Assays were then performed at day 25.

### Immunocytochemistry

*NGN2* neurons were cultured on cover slides coated with poly-L-ornithine/laminin. Cells were fixed in 4% paraformaldehyde in PBS for 15 min. Following permeabilization in PBS containing 0.1% Triton-X100 for 5 min, the slides were blocked with blocking solution (PBS containing 5% normal goat serum) for 1 hour at room temperature. Primary antibodies against MAP-2 (1:500, Abcam, Cat# ab5392) and Beta III Tubulin (1:2000, Promega, Cat# G7121) in blocking solution were added and incubated overnight at 4°C. Cells were then washed with PBS and incubated with Alexa Fluor®-conjugated secondary antibodies (1:500) for 1 hour at room temperature. The following secondary antibodies were used: goat anti-chicken (ThermoFisher Scientific, Cat# A32933) and goat anti-mouse (Abcam, Cat# ab150113). Images were obtained using a confocal microscope system (Zeiss, LSM800).

### Cytokine stimulations of *NGN2* neurons and *C4A*/*C4B* RNA expression

After 25 days, mature *NGN2* neurons were treated with either 1 ng/mLinterleukin (IL)-1β (Thermo Fisher Scientific, cat# PHC0815) or 1 ng/mL IL-6 (Thermo Fisher Scientific, cat# PHC0063), alone or in combination for 24 hours. All experiments were performed in triplicates or duplicates and repeated twice. Following stimulations of cells, total RNA was extracted using Trizol® Reagent (Invitrogen) according to the manufacturer’s protocol. The quantity and purity of total RNA was measured using a NanoDrop® 1000 spectrophotometer (Thermo Scientific). Complementary DNA was synthesized using High-Capacity RNA-to-DNATM Kit (Applied Biosystems) following manufacturer’s protocol. Quantitative PCR was performed with TaqMan Gene Expression Assays (Applied Biosystems) by using the StepOnePlus® Real-time PCR system (Applied Biosystems). Each 20 μL PCR reaction contained 5 μL of cDNA, 300 nM of forward and reverse primer, 250 nM of probe and 10 μL of TaqMan PCR master mix (Applied Biosystems). Primers for *C4A* and *C4B* reactions were as follows (forward: GCAGGAGACATCTAACTGGCTTCT and reverse: CCGCACCTGCATGCTCCT). Specific probes for C4A (FAM-ACC CCT GTC CAG TGT TAG-MGB) and C4B (FAM-ACC TCT CTC CAG TGA TAC-MGB) were used. Data were normalized to the housekeeping gene *GAPDH* (Assay ID. Hs99999905_m1).

### CSF cytokine analyses

IL-1β and IL-6 were measured as part of the Novex Human Ultrasensitive Cytokine 10-Plex Panel (product number at that time: LHC6004; now distrubted by Invitrogen) on the Luminex immunoassay platform. In contrast to the manual provided with the kit, an additional calibration curve point was included to extend the assay dynamic range in the low concentration range. 50 µl sample was diluted with 50 µl of sample diluent, the final sample dilution factor was 1:2. The assay was processed as recommended by the kit instructions. The measurement was performed on a FM3D instrument (operated under software version 4.0, Luminex, Austin, TX, USA). Data analysis was performed using MasterPlex QT 2010 (V 2.0, Hitachi Software Engineering, San Francisco, CA, USA). Standard curves were generated using weighted (1/y^2^) 5-parameter logistics. The analyte concentrations in the samples were calculated based on the standard curves.

### CSF neuronal pentraxins analyses

Preparation of samples and LC-MS/MS analyses were performed as previously described^17^ using micro-high-performance liquid chromatography-mass-spectrometry (6495 Triple Quadrupole LC/MS system, Agilent Technologies). Briefly, in a flow-rate of 0.3 mL/min, sample injection of 40 µL and a 30 min gradient was employed for separation on a Hypersil Gold reversed phase column (dim. 100 × 2.1 mm, particle size 1.9 µm, Thermo Fisher Scientific). The two mobile phases (A and B) were composed of 0.1% formic acid in water (v/v) and 0.1% formic acid/84% acetonitrile in water (v/v), respectively. Electrospray settings were the following: gas temperature 220°C, gas flow 15 L/min, nebulizer 40 psi, sheath gas temperature 200°C, sheath gas flow L/min, capillary voltage in positive mode 3500 V and, nozzle voltage at 500 V. iFunnel settings were the following in positive mode: high-pressure RF 200 V and low-pressure RF 160 V. Individually optimized collision energies and cell accelerator voltages were used for each peptide. The scheduled multiple reaction monitoring method used a delta retention time of 0.8 minutes for each peptide. Quality control samples, consisting of pooled CSF samples obtained from the Neurochemistry Laboratory at Sahlgrenska University Hospital, Mölndal, Sweden, were injected at regular intervals to monitor the performance of the assay over time. The collection and use are in accordance with the Swedish law on biobanks in healthcare (2002:297). Skyline 20.1 (MacCoss Lab Software) was used for peak visualization and inspection. Peak adjustment was performed if required for optimal peak area calculation. Relative peptide concentrations were obtained by summing all measured fragment peak areas for each peptide and dividing that by the sum of the fragment peak areas of the corresponding IS, followed by multiplication of the amount of IS added per µL of CSF.

### Statistics

Available sample size for the initial analysis in the discovery cohort was deemed adequate based on the data presented by Sekar *et al*^1^. [estimated correlation coefficients of at least 0.5 (*P*<10×10^−12^) for between *C4A* and *C4B* CNs versus corresponding RNA expression in 101 subjects, and odds ratio of 1.4 (*P*=2×10^−5^) comparing *C4A* RNA expression in 35 SZ subjects and 70 HCs]. For group comparisons, CSF C4A and C4B concentrations (or *C4A*/*C4B* RNA expression in the different experimental conditions) were log-transformed and analysed by Brown-Forsythe and Welch ANOVA test and corrected for multiple testing using the two-stage linear step-up procedure of Benjamini, Krieger and Yekutieli which controls for false discovery rate (FDR). Correlation analyses were performed using either Pearson’s or Spearman’s correlation coefficient as indicated and dependent on data distribution and sample size. All reported *P*-values were two-sided. *P*<0.05 was interpreted as significant. False discovery rate correction was used to correct for all multiple testing, where applicable and as indicated.

## Data Availability

All data is available from the corresponding author upon reasonable request.

## Acknowledgements

We thank the patients who contributed to this study. Support from the Swedish National Infrastructure for Biological Mass Spectrometry (BioMS) is gratefully acknowledged. C.M.S is supported by grants from the Swedish Research Council, One Mind Foundation, Marianne and Marcus Wallenberg Foundation, and Erling-Persson Family Foundation. F.O received support from the Swedish Brain Foundation and the Swedish Society for Mecial Reseach. KB is supported by the Swedish Research Council (#2017-00915), the Alzheimer Drug Discovery Foundation (ADDF), USA (#RDAPB-201809-2016615), the Swedish Alzheimer Foundation (#AF-742881), Hjärnfonden, Sweden (#FO2017-0243), the Swedish state under the agreement between the Swedish government and the County Councils, the ALF-agreement (#ALFGBG-715986), the European Union Joint Program for Neurodegenerative Disorders (JPND2019-466-236), the National Institute of Health (NIH), USA, (grant #1R01AG068398-01), and the Alzheimer’s Association 2021 Zenith Award (ZEN-21-848495). H.Z. is a Wallenberg Scholar supported by grants from the Swedish Research Council, the European Research Council, and the Swedish Federal Government under the LUA/ALF agreement. NMI received financial support from the State Ministry of Baden-Wuerttemberg for Economic Affairs, Labour and Tourism. S.E. received financial support from the Swedish Research Council. The St. Göran study is supported by grants from the Swedish Research Council (M.L.), the Swedish Foundation for Strategic Research (M.L.), the Swedish Brain Foundation (M.L.), and the Swedish Federal Government under the LUA/ALF agreement (M.L.). KaSP is supported by grants from the Swedish Federal Government under the LUA/ALF agreement (C.M.S., S.E., and S.C.). All illustrations in Figures 1 and 2 were created with BioRender.com.

## Author contributions

J.Gr. and C.M.S. contributed to the overall design, direction, analysis and reporting of the study. F.O., J.Gr., and C.S.M. designed in vitro experiments. F.0., S.G., and N.K. derived neural cultures and performed the in vitro cytokine assay and qPCR. S.D.S. and R.H.P. provided the NGN2 stable neural progenitor cells for the in vitro experiments. C.P.G. performed the ddPCR for the GRIP cohort. S.M. extracted WGS data from KaSP cohort for *C4A* and *C4B*. J.Gr. imputed predicted *C4A* RNA levels from this data. M.S. and C.M.S supervised the genetic analyses. J.H.L. performed shotgun mass spectrometry and helped to develop the method to detect C4A and C4B in CSF. L.S. helped collect and interpret the PPI and habituation data. L.S., S.E., G.E., S.C., F.P., H.F.B., and C.M.S. are part of the KaSP project and helped recruit and collect CSF. H.F.B. performed the cognitive testing of patients in the KaSP cohort. K.B., E.H., H.Z., M.L., A.P., A.G., K.A. and A.I. are part of the GRIP consortium and were involved in recruiting and collecting CSF and patient history. J.N. and A.B analysed KaSP CSF for neuronal pentraxins. V.C. and J.Go. analysed CSF cytokine levels. All authors discussed the results and implications and commented on the manuscript at various stages.

