## Supplementary Information for "Cerebrospinal fluid concentration of complement component 4A is increased in first-episode schizophrenia"

^1^Department of Physiology and Pharmacology, Karolinska Institutet, Stockholm, Sweden; ^2^The Institute of Neuroscience and Physiology, University of Gothenburg, Gothenburg, Sweden; ^3^Department of Clinical Neuroscience, Karolinska Institutet, ^4^Stockholm Health Care Services, Region Stockholm, Sweden; ^5^Department of Anesthesiology, Sahlgrenska University Hospital, Gothenburg, Sweden; ^6^Novartis Institutes of BioMedical Research, Cambridge, MA, USA; ^7^Psychosis Clinic, Sahlgrenska University Hospital, Mölndal, Sweden; ^8^Department of Psychology, University of Gothenburg, Gothenburg, Sweden; ^9^Department of Molecular Medicine and Surgery, Karolinska Institutet, Stockholm, Sweden; ^10^NMI Natural and Medical Sciences Institute at the University of Tübingen, Reutlingen, Germany; ^11^Clinical Neurochemistry Laboratory, Sahlgrenska University Hospital, Mölndal, Sweden; ^12^Department of Neurodegenerative Disease, UCL Institute of Neurology, Queen Square, London, UK; ^13^UK Dementia Research Institute at UCL, London, UK; ^14^Hong Kong Center for Neurodegenerative Diseases, Hong Kong, China; ^15^Center for Quantitative Health, Center for Genomic Medicine and Department of Psychiatry, Massachusetts General Hospital, Boston, MA, USA; ^16^Department of Neuroscience, Uppsala University, Uppsala, Sweden ^17^Department of Medical Epidemiology and Biostatistics, Karolinska Institutet, Stockholm, Sweden.

**Table of contents**

**Supplementary Figure 1.** Cerebrospinal fluid levels of C4A controlling for the use of an antipsychotic medication (<1 month) in the discovery (KaSP) cohort.

**Supplementary Figure 2.** Cerebrospinal fluid levels of C4A in first-episode psychosis patients controlling for the use of antipsychotic medication in the replication (GRIP) cohort.

**Supplementary Figure 3.** Cerebrospinal fluid C4A protein concentration adjusted for *C4A* copy numbers (CNs) in the GRIP cohort.

**Supplementary Figure 4.** Representative validation spectra and linearity graphs of C4 peptides.

**Supplementary Figure 5.** Distribution of *C4AL, C4AS, C4BL and C4BS* copy numbers (CNs) in the GRIP cohort.

**Supplementary Figure 6.** Cerebrospinal fluid levels of C4A in the combined discovery and replication sample.

**Supplementary Table 1.** Demographic and clinical characteristics of the study participants in the replication sample (KaSP).

**Supplementary Table 2.** Demographic and clinical characteristics of the study participants in the discovery sample (GRIP).

**Supplementary table 3.** Correlations of CSF C4A and CSF C4B concentration and the Positive and Negative Syndrome Scale (PANSS) scores, and performance in the key domains of the MATRICS Consensus Cognitive Battery (MCCB).

**Supplementary table 4.** Correlations of CSF C4A concentration on change in EMG recordings during habituation to pulse alone trials, and, at the interstimulus intervals (ISI).

**Supplementary table 5.** Correlations of CSF C4A and CSF C4B concentration on CSF neuronal pentraxins 1 and 2 as well as the neuronal pentraxin receptor.

**Supplementary Table 6.** Inclusion list of the light and heavy peptides with their theoretical masses (m) and charge states (z).

**Supplementary Figure 1.**

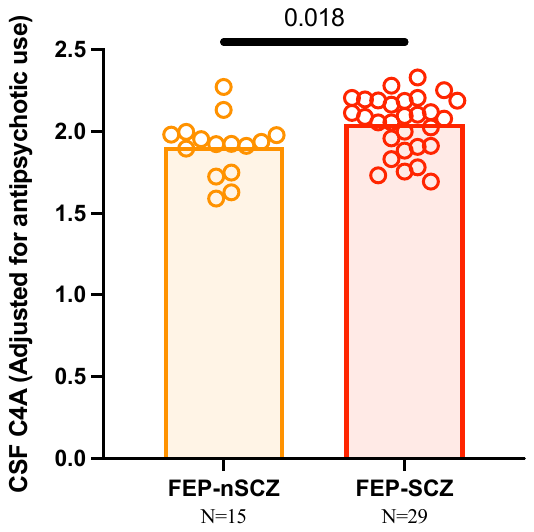

**Supplementary Figure 1. Cerebrospinal fluid levels of C4A after controlling for the use of antipsychotic medication (<1 month) in the discovery (KaSP) cohort.** First episode psychosis subjects who developed schizophrenia (FEP-SCZ) displayed significantly elevated cerebrospinal fluid (CSF) levels of C4A as compared to FEP patients who did not develop SCZ (FEP-nSCZ). CSF C4A data was log-transformed and adjustment for use of antipsychotic medication was performed by using residuals from a linear regression model and then data was analysed using Welch’s *t*-test. The reported p-value is two-sided.

**Supplementary Figure 2**

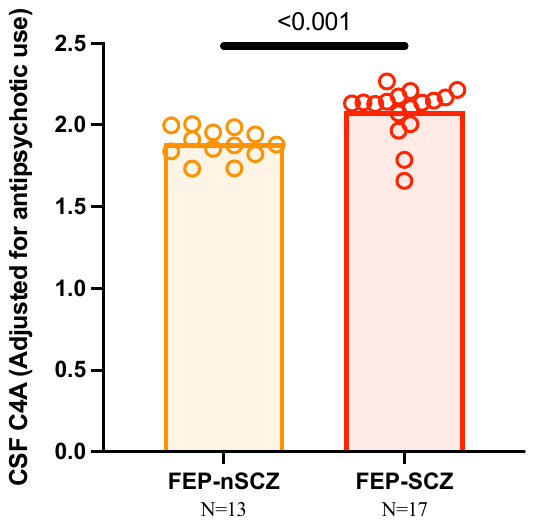

**Supplementary Figure 2. Cerebrospinal fluid levels of C4A in first-episode psychosis patients controlling for the use of antipsychotic medication in the replication (GRIP) cohort.** First episode psychosis subjects who developed schizophrenia (FEP-SCZ) displayed significantly elevated cerebrospinal fluid (CSF) levels of C4A as compared to FEP patients who did not develop SCZ (FEP-nSCZ). CSF C4A data was log-transformed and adjustment for use of antipsychotic medication was performed by using residuals from a linear regression model and then data was analysed using Welch’s *t*-test. The reported p-value is two-sided.

**Supplementary Figure 3.**

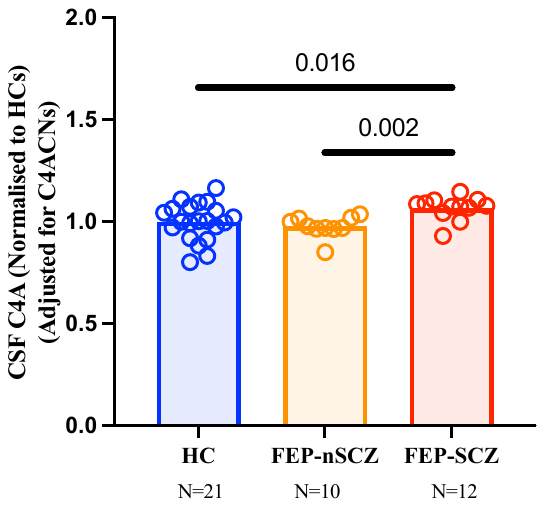

**Supplementary Figure 3. Cerebrospinal fluid C4A protein concentration adjusted for *C4A* Copy numbers (CNs) in the GRIP cohort.** Cerebrospinal fluid (CSF) C4A concentrations (per C4A CNs as measured by droplet digital PCR) were significantly increased in first-episode psychosis patients who developed schizophrenia (FEP-SCZ) as compared to healthy controls (HCs) or FEP patients who did not develop SCZ (FEP-nSCZ). CSF data was log-transformed and adjustment for copy numbers was performed by using residuals from a linear regression model. Data was then analysed by Brown-Forsythe and Welch ANOVA test and corrected for multiple testing using false discovery rate). Plotted data were normalized to mean of HCs. All reported p-values are two-sided.

**Supplementary Figure 4.**

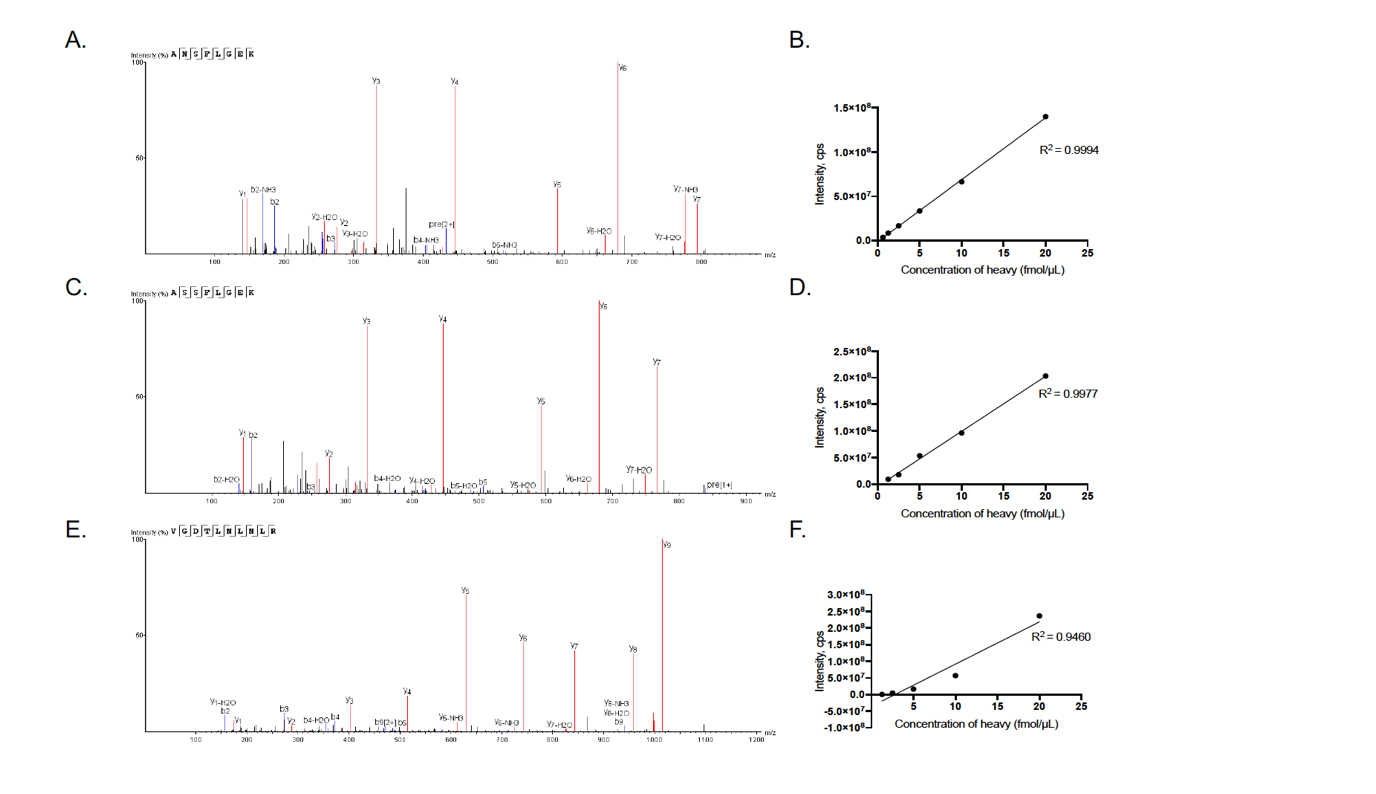

**Supplementary Figure 4:** Representative validation spectra and linearity graphs for C4 peptides. Validation spectra for the unique peptides used to detect C4A (**A.**), C4B (**C.**) and total C4 (tC4) (**E.**) in human cerebrospinal fluid (CSF). **B, D, F** display linear range of heavy peptides in pooled CSF digest for C4A, C4B, and tC4, respectively.

**Supplementary Figure 5.**

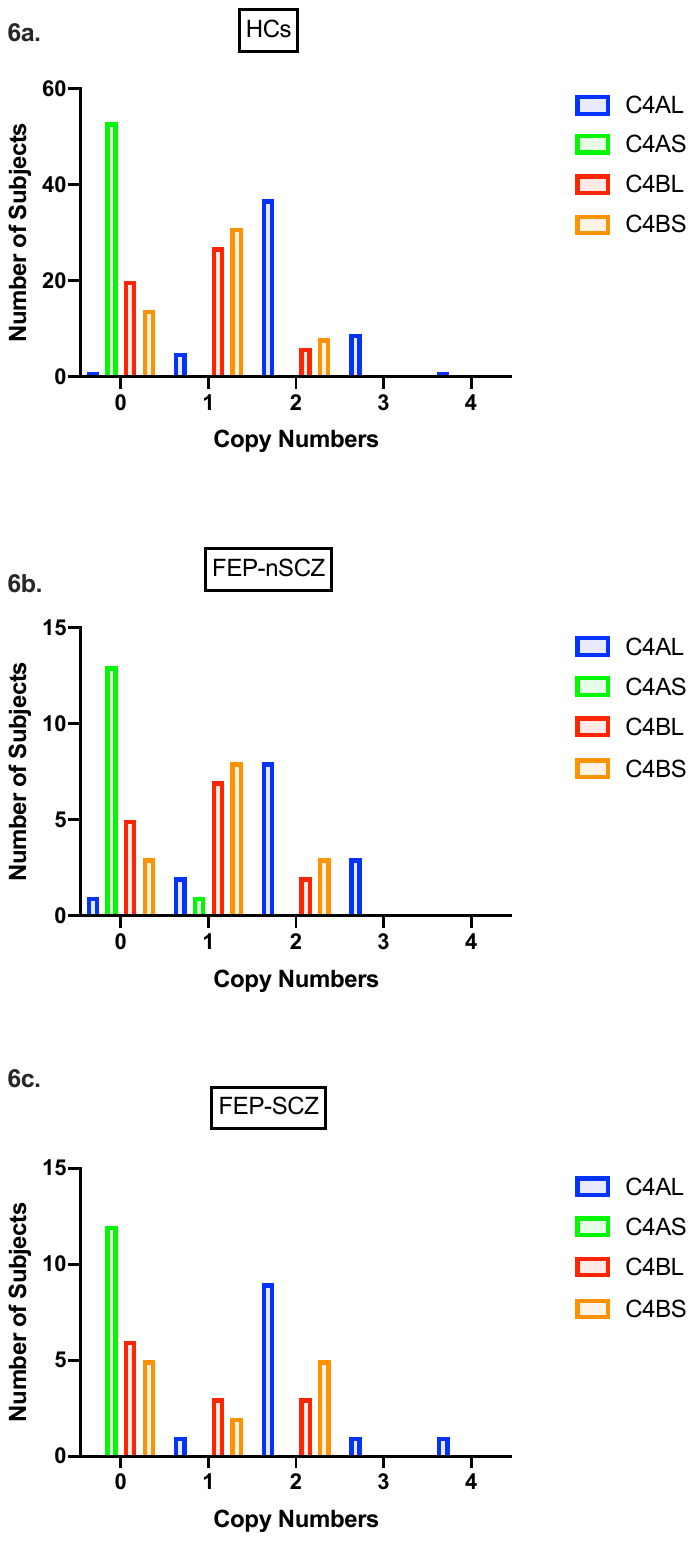

**Supplementary Figure 5. Distribution of *C4AL, C4AS, C4BL and C4BS* copy numbers (CNs) in the GRIP cohort.** Distribution of *C4AL*, *C4AS*, *C4BL* and *C4BS* in 21 healthy controls (HCs; **a**), 11 first episode psychosis subjects who did not develop schizophrenia (FEP-nSCZ), and 12 first-episode psychosis subjects who developed schizophrenia (FEP-SCZ; **c**).

**Supplementary Figure 6.**

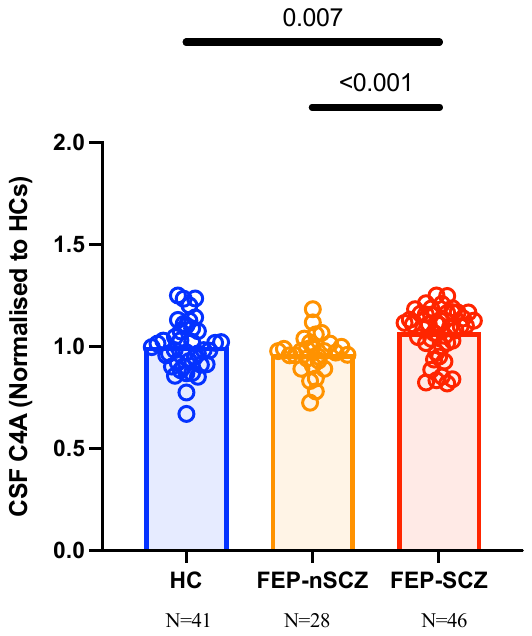

**Supplementary Figure 6. Cerebrospinal fluid levels of C4A in the combined discovery and replication sample.** Cerebrospinal fluid (CSF) C4A concentrations were significantly increased in first-episode psychosis patients who developed schizophrenia (FEP-SCZ; n=46) as compared to healthy controls (HCs; n=41) or FEP patients who did not develop SCZ (FEP-nSCZ; n=28). All data were log-transformed and analysed by Brown-Forsythe and Welch ANOVA test and corrected for multiple testing using False discovery rate). Plotted data were normalized to mean of HCs. All reported p-values are two-sided.

**Supplementary Table 1. Demographic and clinical characteristics of the study participants in the replication sample (KaSP).**

|  | HCs | FEP-nSCZ | FEP-SCZ | P values^1^ |
| --- | --- | --- | --- | --- |
|  | N= 20  Mean | N= 15  Mean | N= 29  Mean | HC vs FEP-nSZ/  HC vs FEP-SZ/  FEP-nSZ vs FEP-SZ |
| Age | 27 | 27 | 30 | 0.978/0.311/0.311 |
| Sex (males/females) | 12/8 | 10/5 | 18/11 | 0.7372/>0.999/>0.999 |
| BMI^2^ | 24^3^ | 23 | 23 | 0.950/0.950/0.950 |
| CSF/Serum Alb ratio^4^ | 4.8 | 7.6^5^ | 6.1^6^ | 0.538/0.433/0.658 |
| Antipsychotics (Y/N) | N/A | 8/7 | 14/15 | >0.999 |
| Nicotine use (Y/N) | 2/18 | 4/11 | 10/19 | 0.367/0.738/0.089 |
| Clinical Global Impression (CGI) | NA | 4.1 | 4.6 | 0.139 |
| Global Assessment of Functioning (GAF) | NA | 33 | 34 | 0.861 |

^1^Healthy controls (HCs), first episode psychosis patients that develop schizophrenia (FEP-SCZ) and first episode psychosis that do not develop schizophrenia (FEP-nSCZ) in the KaSP cohort. Age, BMI and CSF/serum albumin ratio analysed by Brown-Forsythe and Welch ANOVA test and corrected for multiple testing using two-stage linear step-up procedure of Benjamini, Krieger and Yekutieli . Sex, antipsychotics, tobacco use, CGI and GAF were analysed by Fisher’s exact test.

^2^BodyMass Index

^3^Data missing for 1 subject

^4^Cerebrospinal fluid (CSF)/Serum albumin ratio

^5^Data missing for 1 subject

^6^Data missing for 3 subjects

**Supplementary Table 2. Demographic and clinical characteristics of the study participants in the replication cohort (GRIP).**

|  | HCs | FEP-nSCZ | FEP-SCZ | P values^1^ |
| --- | --- | --- | --- | --- |
|  | N=21  Mean | N=14  Mean | N=17  Mean | HC vs FEP-nSCZ/  HC vs FEP-SCZ/  FEP-SCZ vs FEP-nSCZ |
| Age | 35 | 33 | 32 | 0.518/0.415/0.847 |
| Sex (males/females) | 16/5 | 12/2 | 15/2 | 0.676/0.427/>0.999 |
| BMI^2^ | 25 | 26 | 27^3^ | 0.465/0.212/0.497 |
| CSF/Serum Alb ratio^4^ | 4.9 | 4.7^5^ | 5.3 | 0.784/0.784/0.784 |
| Antipsychotics (Y/N) | n/a | 9/5 | 13/4 | 0.693 |
| Tobacco use (Y/N) | 5/16 | 9/4^6^ | 6/11 | 0.014/0.4910/0.139 |
| Relative history of psychiatric illness (Y/N) | NA | 8/4^7^ | 12/2^8^ | 0.239 |
| Sickness benefit (Y/N) | NA | 4/10 | 7/10 | 0.707 |

^1^Healthy controls (HCs), first episode psychosis patients that develop schizophrenia (FEP-SCZ) and first episode psychosis that do not develop schizophrenia (FEP-nSCZ) in the GRIP cohort. Age, BMI and CSF/serum albumin ratio analysed by Brown-Forsythe and Welch ANOVA test and corrected for multiple testing using Two-stage linear step-up procedure of Benjamini, Krieger and Yekutieli False discovery rate (FDR). Sex, antipsychotic and tobacco use, relative history of psychiatric illness and sickness benefit analysed by Fisher’s exact test.

^2^BodyMass Index

^3^Data missing for 1 subject

^4^Cerebrospinal fluid (CSF)/Serum albumin ratio

^5^Data missing for 2 subjects

^6^Data missing for 1 subject

^7^Data missing for 2 subjects

^8^Data missing for 3 subjects

**Supplementary Table 3. Correlations of CSF C4A and CSF C4B concentration and the Positive and Negative Syndrome Scale (PANSS) scores, and performance in the key domains of the MATRICS Consensus Cognitive Battery (MCCB).** All cerebrospinal fluid (CSF) concentrations underwent log transformation and Pearson’s correlation coefficients were used. All *P* values are two sided and correction for multiple testing was performed using Benjamini-Hochberg false discovery rate correction.

|  |  | C4A | | C4B | |
| --- | --- | --- | --- | --- | --- |
|  |  | FEP-nSCZ | FEP-SCZ | FEP-nSCZ | FEP-SCZ |
| Positive and Negative Symptom Scale; PANSS | | | | | |
| Positive sub-scale | r_p_ | 0.04 | -0.25 | -0.26 | 0.09 |
|  | *P* | 0.990 | 0.359 | 0.528 | 0.849 |
|  | N | 15 | 29 | 15 | 28 |
| Negative sub-scale | r_p_ | < -0.01 | -0.23 | -0.17 | -0.04 |
|  | *P* | 0.990 | 0.359 | 0.549 | 0.849 |
|  | N | 15 | 29 | 15 | 28 |
| General sub-scale | r_p_ | 0.11 | -0.06 | -0.53 | 0.25 |
|  | *P* | 0.990 | 0.775 | 0.129 | 0.618 |
|  | N | 15 | 29 | 15 | 28 |
|  |  | C4A | | C4B | |
| MATRICS Consensus Cognitive Battery (MCCB) | | | | | |
| TMT | r_p_ | -0.53 | 0.36 | -0.36 | -0.07 |
|  | *P* | 0.253 | 0.460 | 0.776 | 0.997 |
|  | N | 15 | 27 | 15 | 26 |
| BACS-SC | r_p_ | 0.51 | 0.33 | 0.32 | 0.09 |
|  | *P* | 0.253 | 0.460 | 0.776 | 0.997 |
|  | N | 15 | 27 | 15 | 26 |
| HVLT-R | r_p_ | 0.47 | 0.25 | 0.27 | 0.06 |
|  | *P* | 0.253 | 0.606 | 0.776 | 0.997 |
|  | N | 15 | 25 | 15 | 26 |
| WMS III SS | r_p_ | 0.36 | 0.22 | 0.23 | 0.04 |
|  | *P* | 0.463 | 0.606 | 0.776 | 0.997 |
|  | N | 15 | 27 | 15 | 26 |
| LNS | r_p_ | 0.26 | 0.21 | 0.19 | 0.05 |
|  | *P* | 0.686 | 0.606 | 0.776 | 0.997 |
|  | N | 15 | 27 | 15 | 26 |
| NAB-MAZES | r_p_ | -0.19 | 0.16 | -0.18 | -0.03 |
|  | *P* | 0.733 | 0.690 | 0.776 | 0.997 |
|  | N | 15 | 27 | 15 | 26 |
| BVMT-R | r_p_ | 0.17 | 0.05 | -0.14 | -0.03 |
|  | *P* | 0.733 | 0.978 | 0.776 | 0.997 |
|  | N | 15 | 27 | 15 | 26 |
| Fluency | r_p_ | 0.13 | 0.03 | 0.14 | 0.21 |
|  | *P* | 0.733 | 0.978 | 0.776 | 0.997 |
|  | N | 15 | 27 | 15 | 26 |
| MSCEIT ME | r_p_ | 0.09 | 0.02 | 0.11 | -0.25 |
|  | *P* | 0.733 | 0.978 | 0.776 | 0.997 |
|  | N | 15 | 27 | 15 | 26 |
| CPT-IT | r_p_ | -0.09 | <0.01 | 0.05 | <-0.01 |
|  | *P* | 0.733 | 0.978 | 0.866 | 0.997 |
|  | N | 15 | 27 | 15 | 25 |

**Supplementary table 4. Correlations of CSF C4A concentration with change in EMG recordings during habituation to pulse alone trials, and, at the interstimulus interval (ISI) between the prepulse and the pulse at 30, 60, and 120 ms.** All C4A cerebrospinal fluid (CSF) concentrations underwent log transformations and Pearson’s correlation coefficients were used. All *P* values are two sided and correction for multiple testing was performed using Benjamini-Hochberg false discovery rate correction.

| ISI 30 | r_p_ | 0.22 |
| --- | --- | --- |
|  | *P* | 0.483 |
|  | N | 42 |
| ISI 60 | r**_p_** | -0.07 |
|  | *P* | 0.688 |
|  | N | 41 |
| ISI 120 | r**_p_** | 0.12 |
|  | *P* | 0.688 |
|  | N | 41 |
| Habituation 1 | r**_p_** | 0.31 |
|  | *P* | 0.072 |
|  | N | 43 |
| Habituation 2 | r**_p_** | 0.40 |
|  | *P* | 0.031 |
|  | N | 43 |
| Habituation 3 | r**_p_** | 0.43 |
|  | *P* | 0.031 |
|  | N | 43 |
| Habituation 4 | r**_p_** | 0.27 |
|  | *P* | 0.101 |
|  | N | 44 |
| Habituation 5 | r**_p_** | 0.35 |
|  | *P* | 0.044 |
|  | N | 43 |
| Habituation 6 | r**_p_** | 0.40 |
|  | *P* | 0.031 |
|  | N | 42 |
| Habituation 7 | r**_p_** | 0.27 |
|  | *P* | 0.117 |
|  | N | 41 |
| Habituation 8 | r**_p_** | 0.23 |
|  | *P* | 0.160 |
|  | N | 41 |
| Habituation 9 | r**_p_** | 0.37 |
|  | *P* | 0.040 |
|  | N | 42 |
| Habituation 10 | r**_p_** | 0.39 |
|  | *P* | 0.031 |
|  | N | 42 |
| Habituation 11 | r**_p_** | 0.24 |
|  | *P* | 0.139 |
|  | N | 42 |
| Habituation 12 | r**_p_** | 0.18 |
|  | *P* | 0.235 |
|  | N | 43 |
| Habituation 13 | r**_p_** | 0.38 |
|  | *P* | 0.031 |
|  | N | 43 |
| Habituation 14 | r**_p_** | 0.34 |
|  | *P* | 0.044 |
|  | N | 44 |

**Supplementary table 5. Correlations of cerebrospinal fluid (CSF) C4A and C4B protein concentration with CSF neuronal pentraxins 1 and 2, as well as the neuronal pentraxin receptor.** CSF from 19 healthy controls and 43 first episode psychosis patients from the KaSP cohort were analysed, all CSF concentrations underwent log transformations and Pearson’s correlation and adjusted for age, sex and diagnosis. All *P* values are two sided and correction for multiple testing was performed using Benjamini-Hochberg false discovery rate correction.

|  |  | Neuronal pentraxin 2 | Neuronal pentraxin 1 | Neuronal pentraxin receptor |
| --- | --- | --- | --- | --- |
| C4A | r**_p_** | -0.28 | -0.30 | -0.32 |
|  | *P* | 0.033 | 0.033 | 0.033 |
|  | N | 57 | 57 | 57 |
| C4B | r**_p_** | 0.24 | 0.07 | 0.07 |
|  | *P* | 0.213 | 0.622 | 0.622 |
|  | N | 56 | 56 | 56 |

**Supplementary Table 6.** Inclusion list of the light and heavy peptides with their theoretical masses (m) and charge states (z).

| **Proteins^a^** | **Peptide Sequence^b^** | **m/z^c^** | **z** |
| --- | --- | --- | --- |
| C4 total | VGDTLNLNLR | 557.81 | 2 |
|  | VGDTLNLNL**R** | 562.82 | 2 |
| CO4A | ANSFLGEK | 433.22 | 2 |
|  | ANSFLGE**K** | 437.23 | 2 |
| CO4B | ASSFLGEK | 419.71 | 2 |
|  | ASSFLGE**K** | 423.72 | 2 |

**^a^**	C4 total is a peptide representing the total amounts of Complement component C4 (peptide common for isotypes C4A and C4B).

CO4A and CO4B are unique peptides for Complement C4A and C4B, respectively.

**^b^**	The bold amino acids are modified.

**^c^** Mass over charge ratio for the peptides.
